# Catheter Ablation Approach Targeting Epicardial Connections to the Right Pulmonary Vein Antrum Detected before Pulmonary Vein Isolation

**DOI:** 10.1101/2024.03.04.24303750

**Authors:** Yosuke Nakatani, Yutaka Take, Shingo Yoshimura, Ryoya Takizawa, Koji Goto, Kenichi Kaseno, Yumiko Haraguchi, Koki Kimura, Takehito Sasaki, Yuko Miki, Kohki Nakamura, Shigeto Naito

## Abstract

**Background:** Epicardial connections from surrounding structures to the right pulmonary vein (PV) antrum impede the PV isolation. This study aimed to evaluate the efficacy of an ablation approach targeting epicardial connections for right PV isolations.

**Methods:** We prospectively enrolled 124 atrial fibrillation patients who underwent initial PV isolations. We identified the activation breakthrough site into the right PV antrum (BT-RPV) on the activation map created during high right atrial pacing before the PV isolation. BT-RPV sites were targeted when right PV isolations were not achieved by a wide antral circumferential ablation (WACA).

**Results:** A BT-RPV was observed in 83 cases (67%). BT-RPV sites were predominantly located on the anterior part of the carina (45% of BT-RPV sites). PV isolation was achieved by a WACA in all 41 cases without BT-RPVs. Among the cases with BT-RPVs, the PV isolation was achieved by a WACA in all 48 cases where all BT-RPV sites were covered by the PV isolation line. Conversely, the PV isolation was completed by a WACA in only 5 out of 35 cases (14%) when not all BT-RPV sites were covered. In 30 cases where the WACA did not achieve the PV isolation, 35 sites were targeted for the BT-RPV ablation. The initial BT-RPV ablation led to a PV isolation at 20 sites, while the remaining 15 BT-RPV sites required a repeat BT-RPV ablation. The ablated area of a successful BT-RPV ablation was 0.9 [0.6–1.2] cm^2^, corresponding to the area activated within 15 [14-16] ms after the BT-RPV emergence. Ablating the area that was activated within 14 ms after the BT-RPV emergence was associated with successful PV isolations (sensitivity 91% and specificity 100%).

**Conclusion:** Ablation targeting BT-RPV sites is effective for a right PV isolation. However, an extensive ablation area is required to eliminate BT-RPVs.

**CLINICAL PERSPECTIVE:** *What is Known?:*

- Pulmonary vein isolation using a wide antral circumferential ablation is sometimes complicated by epicardial connections from neighboring structures to the right pulmonary vein antrum.
- The intercaval muscular fibers and septopulmonary bundle can act as substrates for epicardial conduction pathways.

*What the Study Adds:*

- Epicardial connections to the right pulmonary vein antrum have been identified in 67% of patients with atrial fibrillation.
- Targeted ablation at activation breakthrough sites can effectively disrupt epicardial connections to the right pulmonary vein antrum, significantly enhancing the rate of atrial fibrillation-free survival.
- For the complete interruption of these epicardial pathways, ablation is necessary in a specific region, indicated by the area activated within a certain time frame following the onset of the epicardial activation in the right pulmonary vein antrum.

**Tweet:** Ablation targeting the breakthrough point of epicardial connections to the right pulmonary vein antrum is effective for pulmonary vein isolation.

**Graphic abstract:** 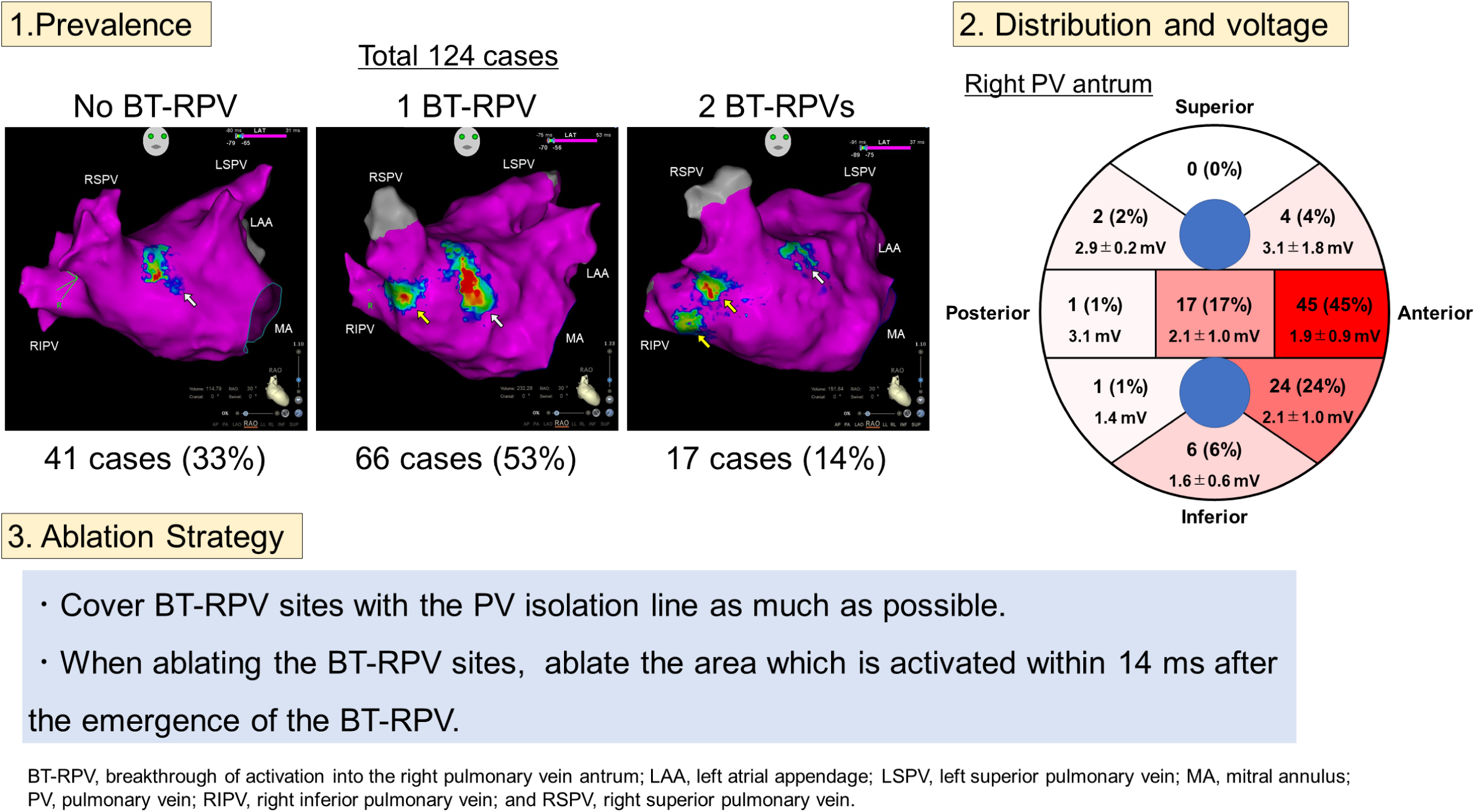

## INTRODUCTION

The process of interatrial conduction is primarily facilitated through three main pathways: Bachmann’s bundle, the fossa ovalis, and the coronary sinus musculature.^1–3^ However, previous autopsy studies^4–5^ have revealed additional muscular fibers that connect between the right and left atria, which may provide epicardial substrates for posterior interatrial electrical conduction.^3,6–10^ Furthermore, there exists the possibility that the rightward branch of the septopulmonary bundle may extend towards the carina of the right pulmonary veins (PVs), potentially serving as a substrate for epicardial conduction.^8,11,12^

A PV isolation with a wide antral circumferential ablation (WACA) has become a cornerstone approach for catheter ablation of atrial fibrillation (AF).^13^ However, in some cases, a circumferential ablation of the antrum alone does not complete the PV isolation, and requires ablation inside the PV isolation line. In those cases, electrical excitation through epicardial pathways may hinder the PV isolation, suggesting that intentional ablation targeting those epicardial pathway connections could be effective for the PV isolation. While the efficacy of such approaches has been suggested in retrospective studies,^6–8^ there have been few prospective studies evaluating their effectiveness.

In this study, we prospectively evaluated the effectiveness of a systematic ablation approach that involved assessing the epicardial connections to the right PV antrum prior to ablation and subsequently targeting them for ablation.

## METHODS

### Study population

This study was a prospective evaluation of consecutive patients with AF who underwent a PV isolation at the Gunma Prefectural Cardiovascular Center (Maebashi, Japan) from November 2021 to May 2023. We included all types of AF, including paroxysmal AF (lasting <7 days), persistent AF (lasting ≥7 days but <1 year), and long-standing persistent AF (lasting ≥1 year). We excluded patients where AF was not terminated before the PV isolation and those with a prior catheter ablation. The study protocol was approved by the Research and Ethics Committee of the institution and conducted in accordance with the principles of the Declaration of Helsinki. All patients provided written informed consent as per its institutional guidelines.

### Ablation Protocol

All antiarrhythmic drugs were discontinued at least five half-lives before the catheter ablation. No patients received oral amiodarone. All procedures were performed under conscious sedation with propofol or dexmedetomidine. A temperature monitoring probe (SensiTherm, Abbott, St. Paul, MN) was placed in the esophagus. Sheath introducers were inserted through the right femoral vein. A steerable decapolar catheter was placed in the coronary sinus. The transseptal puncture was performed using fluoroscopic landmarks or intracardiac echocardiography, and two 8-F SL0 sheaths or an 8-F SL0 sheath and a steerable sheath (AGILIS, Abbott) were advanced into the left atrium.

Electroanatomical mapping was performed using a 3D mapping system (CARTO 3 system, Biosense-Webster, Diamond Bar, CA). When AF persisted at the timing of the mapping, external cardioversion was performed. During high right atrial pacing with a pacing cycle length of 600 ms, sequential contact mapping of the left atrium was performed with a high-density mapping catheter (PENTARAY or OCTARAY; Biosense-Webster). The contact of the mapping catheter was validated by stable electrograms, the distance to the geometry surface, and catheter motion concordant with the cardiac silhouettes assessed by fluoroscopy. Each point was collected using the automated CONFIDENSE mapping module (Biosense-Webster). The initial CONFIDENSE settings included the following: the cycle length stability, which measured the cycle length between two beats and only accepted those within a 10% range of the cycle length; electrode position stability, which only accepted points collected less than 2 mm away from the previous beat; local activation time stability, which rejected points with a local activation time of more than 3 ms compared with that of the previous beat; tissue proximity filter, which used impedance measurements to determine the proximity of electrode to the cardiac surface; and density, which rejected points within 1 mm from the previous points. A fill/color interpolation threshold of 15 mm was used.

Breakthrough of the activation into the right PV antrum (BT-RPV) was assessed in the activation map of the left atrium. BT-RPVs were defined as a centrifugal activation occurring in the right PV antrum, which preceded the activations coming to the area through other interatrial conduction pathways, such as Bachmann’s bundle, the foramen ovale, and the coronary sinus. Representative cases are shown in Fig. 1. BT-RPV sites were tagged on the map of the left atrium.

**Figure 1.**
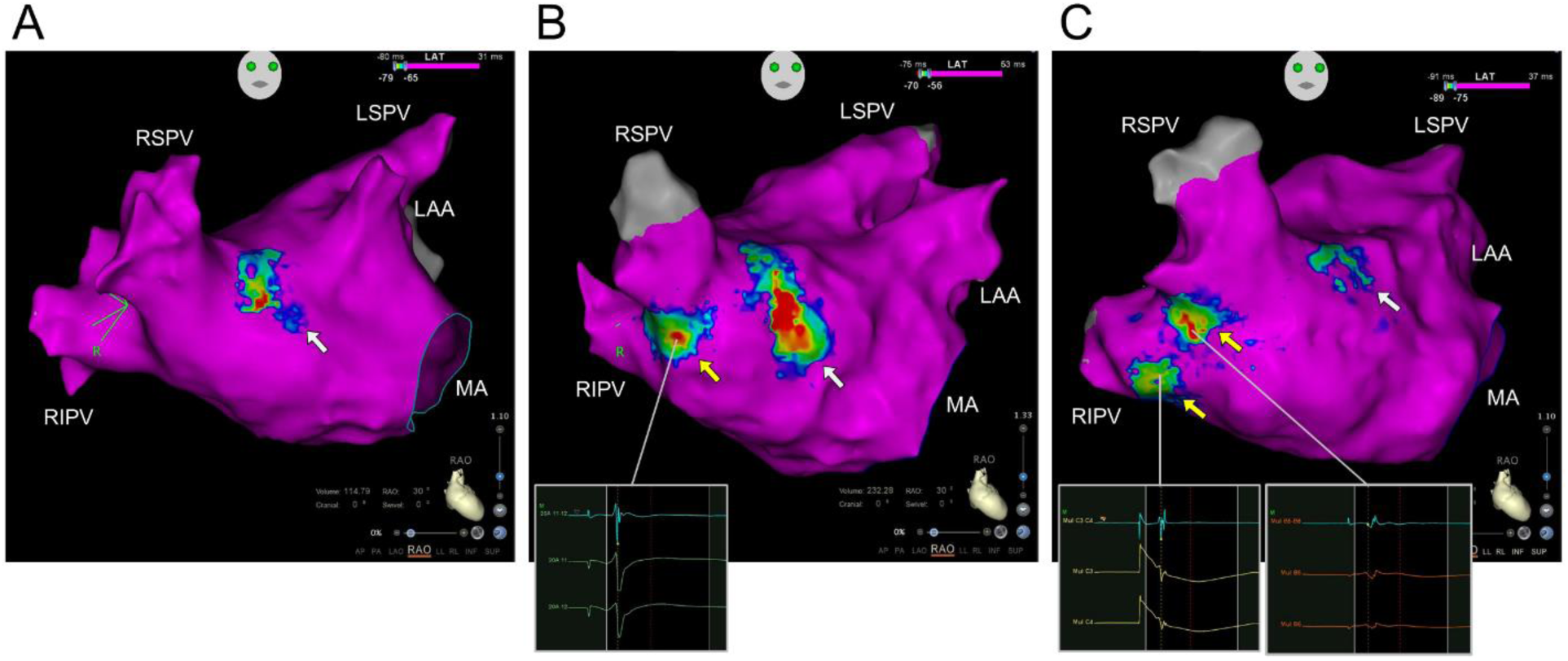
Representative cases of the activation map of the left atrium created during high right atrial pacing. (A) A case illustrating the absence of a breakthrough of the activation into the right pulmonary vein antrum (BT-RPV). Activation is solely via Bachmann’s bundle, indicated by the white arrow. (B) A case with a single BT-RPV site. The BT-RPV, marked with the yellow arrow, is observed on the anterior part of the carina. (C) A case with two distinct BT-RPV sites. These sites are located on the anterior part of the carina and the right inferior pulmonary vein (RIPV). Fragmented potentials were observed on the BT-RPV sites. LAA indicates left atrial appendage; LSPV, left superior pulmonary vein; MA, mitral annulus; and RSPV, right superior pulmonary vein.

The PV isolation was performed with a WACA under the guidance of a 3D computed tomographic image of the left atrium integrated by the CARTO Merge system (Biosense Webster). An irrigated tip radiofrequency catheter (ThermoCool SmartTouch SF, Biosense-Webster, Inc.) with a contact force sensor was used for the radiofrequency application. A power setting of 40 W and target ablation index of 550 were utilized for the PV isolation. In areas near the esophagus, a power setting of 25-35 W and duration of 30 s were utilized, and the radiofrequency application was ceased when the esophageal temperature exceeded 41 °C. During the right PV isolation procedure, ablation lesions were created to pass through BT-RPV sites as much as possible. Circular catheters placed within the PVs were used to confirm the entrance block, by the elimination or dissociation of the recorded PV potentials, and the exit block, by pacing from the catheters.

The right PV isolation was performed first. When the right PV isolation was not achieved by the first-pass ablation, ablation targeting BT-RPV sites was performed in cases with BT-RPV sites that were not covered by the WACA line. If the right PV isolation was achieved during ablation of the BT-RPV sites, adjunctive ablation was performed in the area around the BT-RPV site. If the right PV isolation could not be achieved by the BT-RPV ablation, we searched for gaps on the PV isolation line, and ablation was performed at the gaps if they were found. In cases where no gap was found or ablation on the gaps was not effective, a repeat BT-RPV ablation was performed. In the repeat BT-RPV ablation, a more extensive area around the BT-RPV was ablated than in the first BT-RPV ablation. The left PV isolation was performed after completing the right PV isolation. Following the completion of the left PV isolation, a reconnection of the right PV isolation was assessed. Additionally, under an isoproterenol infusion, 10 mg of adenosine triphosphate was intravenously administered to reveal any potential reconnections. When a PV reconnection was observed in the right PV isolation, a repeat BT-RPV ablation was performed. When reconnections and dormant conduction were no longer observed on the bilateral PV isolation lines, we created a cavotricuspid isthmus line and finished the procedure.

### Offline assessment of the BT-RPV sites

To define the location of the BT-RPV sites, we divided the right PV antrum into 9 parts: the anterior, superior, and posterior parts of the right superior PV antrum, the anterior, middle, and posterior parts of the carina, and the anterior, inferior, and posterior parts of the right inferior PV antrum. Moreover, the bipolar voltage of the local electrograms at the BT-RPV sites was measured on the voltage map.

We assumed that connections of an epicardial pathway to the right PV antrum extended over a certain area, and it would be necessary to ablate the entire area of the connections. Therefore, to investigate the appropriate ablation area for the BT-RPV ablation, we compared the lesion size between the successful and unsuccessful BT-RPV ablation sites. The lesion size was measured by manually tracing the area with ablation tags. Furthermore, when the epicardial pathway connects across a certain extent of the area, this area should be activated within a short period from the emergence of the BT-RPV. Therefore, we assessed the time between the emergence of the BT-RPV and the activation of the area that needed to be ablated to eliminate the BT-RPV. In the local activation time color bar of the activation map, we set the first marker at the timing of the BT-RPV emergence and moved the second marker to adjust the highlighted area (Fig. 2). That highlighted area represented the area activated within the time between the emergence of the BT-RPV and the timing of the second marker. We adjusted the second marker to make all RT-RPV ablation tags covered by the highlighted area. We measured the time difference between the first marker and the second marker and compared that between the successful and unsuccessful BT-RPV ablation sites.

**Figure 2.**
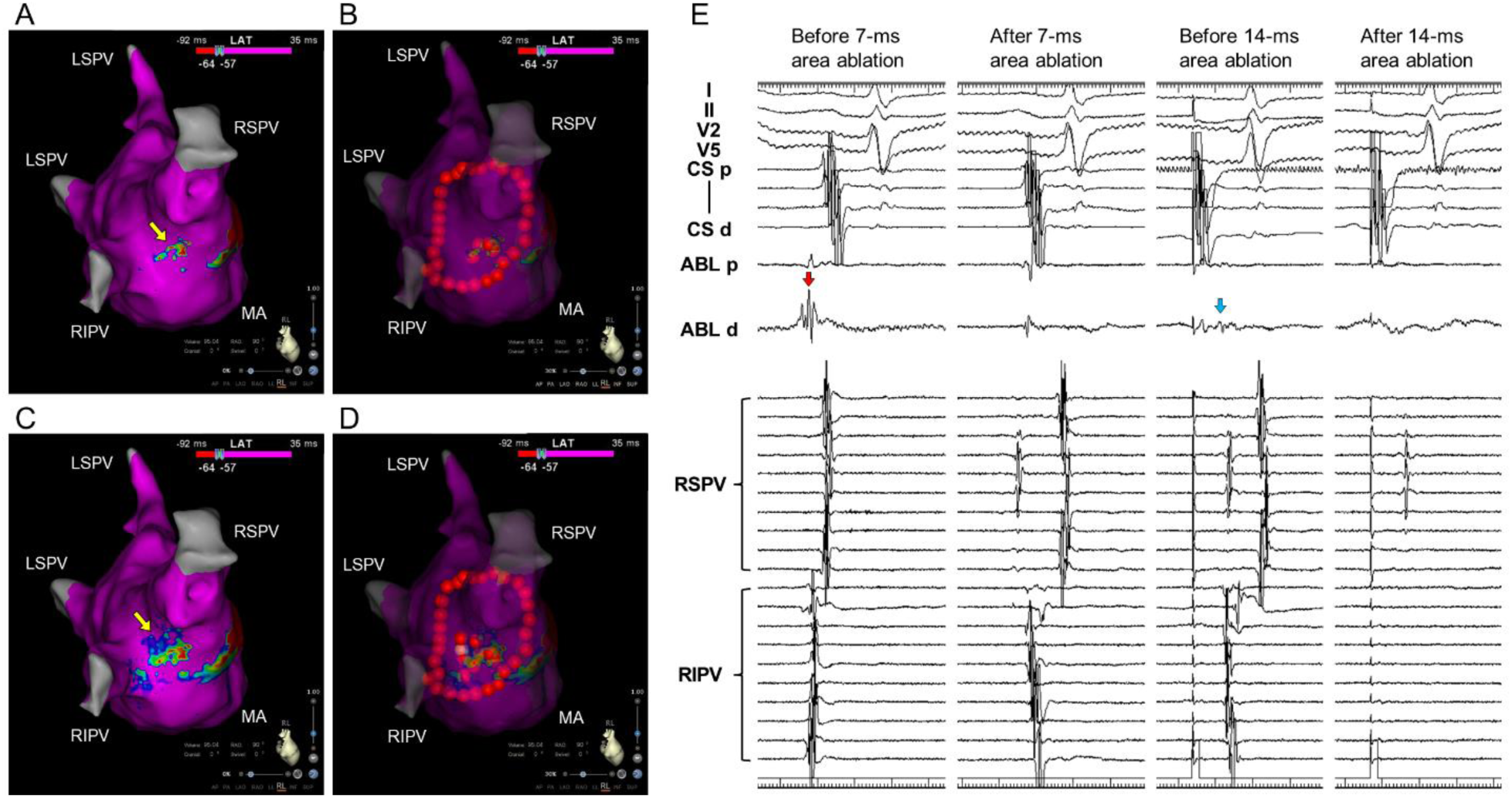
A representative case of ablation targeting the breakthrough of the activation into the right pulmonary vein antrum (BT-RPV). (A) The area activated within 7 ms after the emergence of the BT-RPV (a yellow arrow). (B) Wide antral circumferential ablation and ablation of the area activated within 7 ms. (C) The area activated within 14 ms. (D) Ablation of a BT-RPV and the area activated within 14 ms. (E) Tracings during the BT-RPV ablation. Before the ablation, a fragmented potential was observed at the BT-RPV site (a red arrow). Ablation of the area activated within 7 ms delayed the pulmonary vein potentials. A low voltage potential was observed in the area activated within 14 ms, and ablation at that site completed the pulmonary vein isolation. CS indicates coronary sinus; LIPV, left inferior pulmonary vein. The other abbreviations are as in Fig. 1.

### Post-procedure care and follow-up

A clinical interview and surface 12-lead electrocardiogram were performed the day after ablation and monthly thereafter during visits to the outpatient clinic. Twenty-four-hour Holter monitoring was performed as needed. Documentation of AF, lasting ≥30 s on a surface 12-lead electrocardiogram or Holter monitor, was used to identify AF recurrence. AF occurring in the first 3 months after the ablation (blanking period) was censored.

### Statistical analysis

The Shapiro-Wilk test of normality was used to assess whether quantitative data conformed to the normal distribution. Continuous data are expressed as the mean ± standard deviation when following a normal distribution and as the median [interquartile range Q1–Q3] otherwise. Categorical data are expressed as numbers (%). Independent continuous variables were compared using independent-sample parametric (unpaired Student’s t-test) or non-parametric tests (Mann-Whitney U test) depending on the data normality. Independent categorical variables were compared using the Chi-square test when the expected frequencies were ≥5 and Fisher’s exact test when they were <5. Multiple-group comparisons were performed using a 1-way ANOVA. If significant differences were identified, post hoc Bonferroni-adjusted pairwise comparisons were performed. The receiver-operating characteristic curve analysis was performed to assess the optimal cutoff values. The log-rank test was performed to determine differences in the AF-free survival between patients with and without BT-RPVs. We considered a P <0.050 as statistically significant.

## RESULTS

### Patient characteristics

We conducted a prospective review to assess the eligibility for inclusion in this study among 144 consecutive patients with AF. We excluded 12 patients due to prior catheter ablation, and the other patients underwent the ablation procedure with our protocol. We excluded 8 patients because AF did not terminate before the PV isolation. Ultimately, the analysis consisted of 124 patients in which ablation with our protocol was completed.

The patient characteristics are summarized in Table 1. The median age was 70 [59–74] years, and 73% of the patients were male. The AF type was paroxysmal, persistent, and long-standing persistent in 62%, 33%, and 5% of patients, respectively. The time from the AF onset to the ablation was 5 [3–24] months. Structural heart disease and congestive heart failure were observed in 21% and 10% of patients, respectively. Antiarrhythmic drugs were administered in 31% of the patients before ablation. The left atrial volume and left ventricular ejection fraction were 115 [95–154] ml and 60 [56–65] %, respectively.

**Table 1.** Comparison of the patient characteristics between the patients with and without epicardial connections.

|  | All cases<br>(n = 124) | Epicardial<br>connection (+)<br>(n = 83) | Epicardial<br>connection (-)<br>(n = 41) | P value |
| --- | --- | --- | --- | --- |
| Age, years | 70 [59–74] | 71 [59–74] | 69 [60–75] | 0.875 |
| Male gender | 90 (73) | 58 (70) | 32 (78) | 0.337 |
| AF type |  |  |  | 0.681 |
| Paroxysmal | 77 (62) | 51 (61) | 26 (63) |  |
| Persistent | 41 (33) | 27 (33) | 14 (34) |  |
| Long-standing persistent | 6 (5) | 5 (6) | 1 (2) |  |
| AF onset to ablation | 5 [3–24] | 5 [3–23] | 8 [3–39] | 0.191 |
| Structural heart disease | 26 (21) | 16 (19) | 10 (24) | 0.681 |
| Congestive heart failure | 13 (10) | 10 (12) | 3 (7) | 0.418 |
| Hypertension | 66 (53) | 49 (59) | 17 (41) | 0.065 |
| Diabetes mellitus | 19 (15) | 13 (16) | 6 (15) | 0.881 |
| Past history of stroke | 5 (4) | 4 (5) | 1 (2) | 0.526 |
| CHADS2 score | 1 [0–2] | 1 [0–2] | 1 [0–1] | 0.147 |
| Antiarrhythmic drugs | 38 (31) | 24 (29) | 14 (34) | 0.552 |
| Left atrial volume, cm<sup>2</sup> | 115 [95–154] | 112 [95–155] | 119 [92–152] | 0.848 |
| Left ventricular ejection<br>fraction, % | 60 [56–65] | 60 [95–155] | 60 [60–65] | 0.367 |

Data are the median [interquartile range Q1–Q3] or number (%) of patients. AF, atrial fibrillation.

### Prevalence, distribution, and local voltage of the BT-RPV sites

A median of 7170 [4320–9784] points was acquired during the mapping of the left atrium. BT-RPVs were observed in 83 cases (67%). Of those, 66 cases (53%) exhibited one BT-RPV site, while 17 (14%) had two distinct BT-RPV sites (Fig. 1). Fragmented potentials were generally observed at the BT-RPV sites. The distribution and local voltage of the BT-RPV sites are shown in Fig. 3. The most common location of the BT-RPV sites was the anterior part of the carina (45% of BT-RPV sites), followed by the anterior part of the right inferior PV antrum (24%), and the middle part of the carina (17%). The median local voltage at the BT-RPV sites was 1.9 [1.4–2.6] mV. No statistically significant difference in the local voltage was observed among the various BT-RPV sites (P = 0.232).

**Figure 3.**
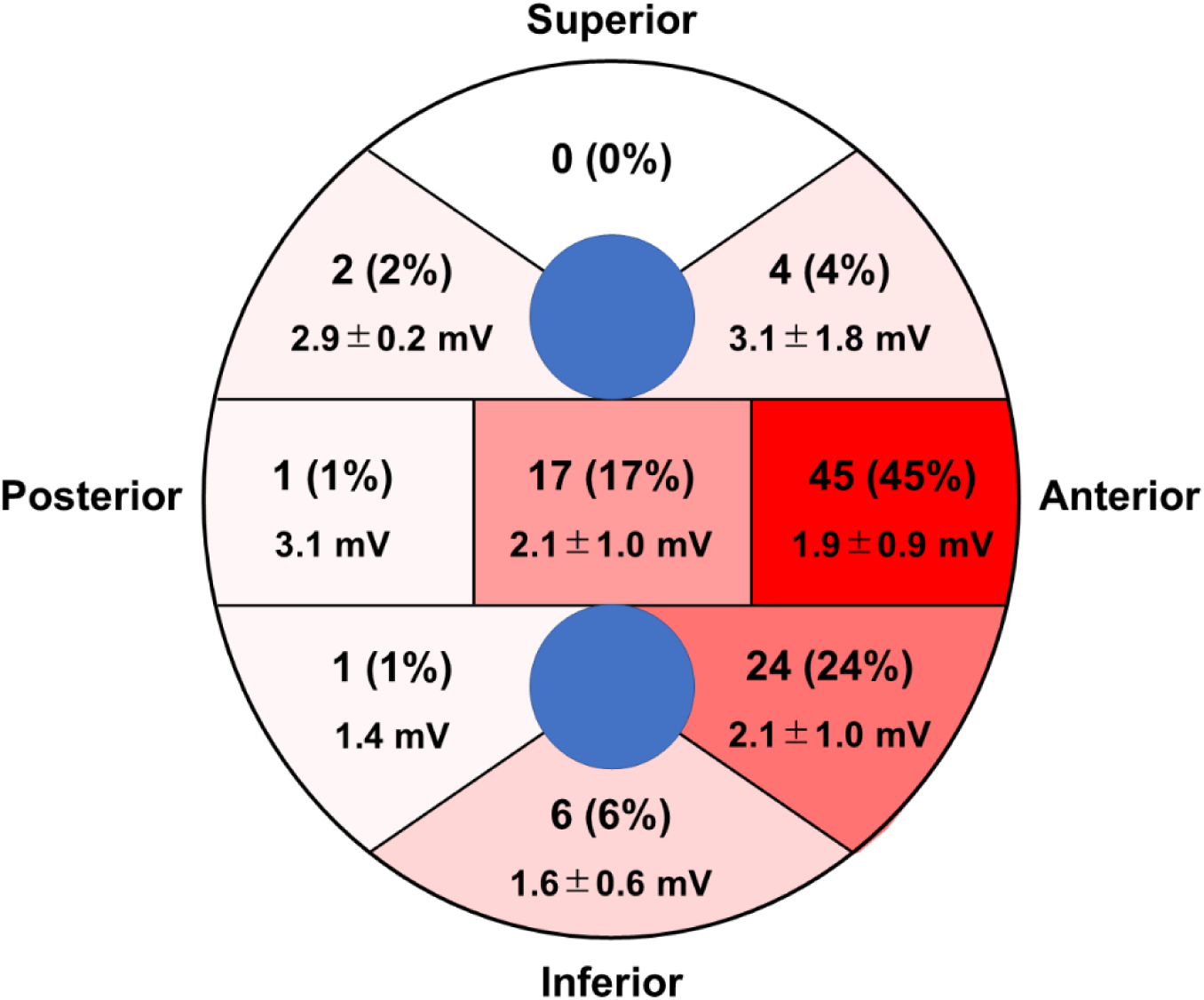
Schematic diagram showing the distribution and local voltage at the breakthrough sites of the activation into the right pulmonary vein antrum (BT-RPV). The schematic diagram represents the right pulmonary vein antrum. The frequency of the BT-RPVs is indicated by the numbers (%), and the local voltage is presented as the mean ± standard deviation. Each area is shaded in progressively darker red according to the frequency of the BT-RPVs.

### Impact of BT-RPVs on the right PV isolation

The results of the WACA for the right PVs are summarized in Fig. 4. Among 41 cases without BT-RPVs, a first-pass ablation completed the PV isolation in 39 cases (95%). In the remaining 2 cases, additional ablation targeting gaps on the PV isolation line was necessary. Among the 83 cases with BT-RPVs, the PV isolation line passed all BT-RPV sites in 48 cases. Of those 48 patients, a first-pass ablation was successful in 46 cases (96%), and the remaining 2 required an additional line-gap ablation. Conversely, among 35 cases in whom the PV isolation line did not cover all BT-RPV sites, a first-pass ablation was only effective in 5 cases (14%). In the other 30 cases in which the first-pass ablation was insufficient, ablation targeting the BT-RPV sites was performed.

**Figure 4.**
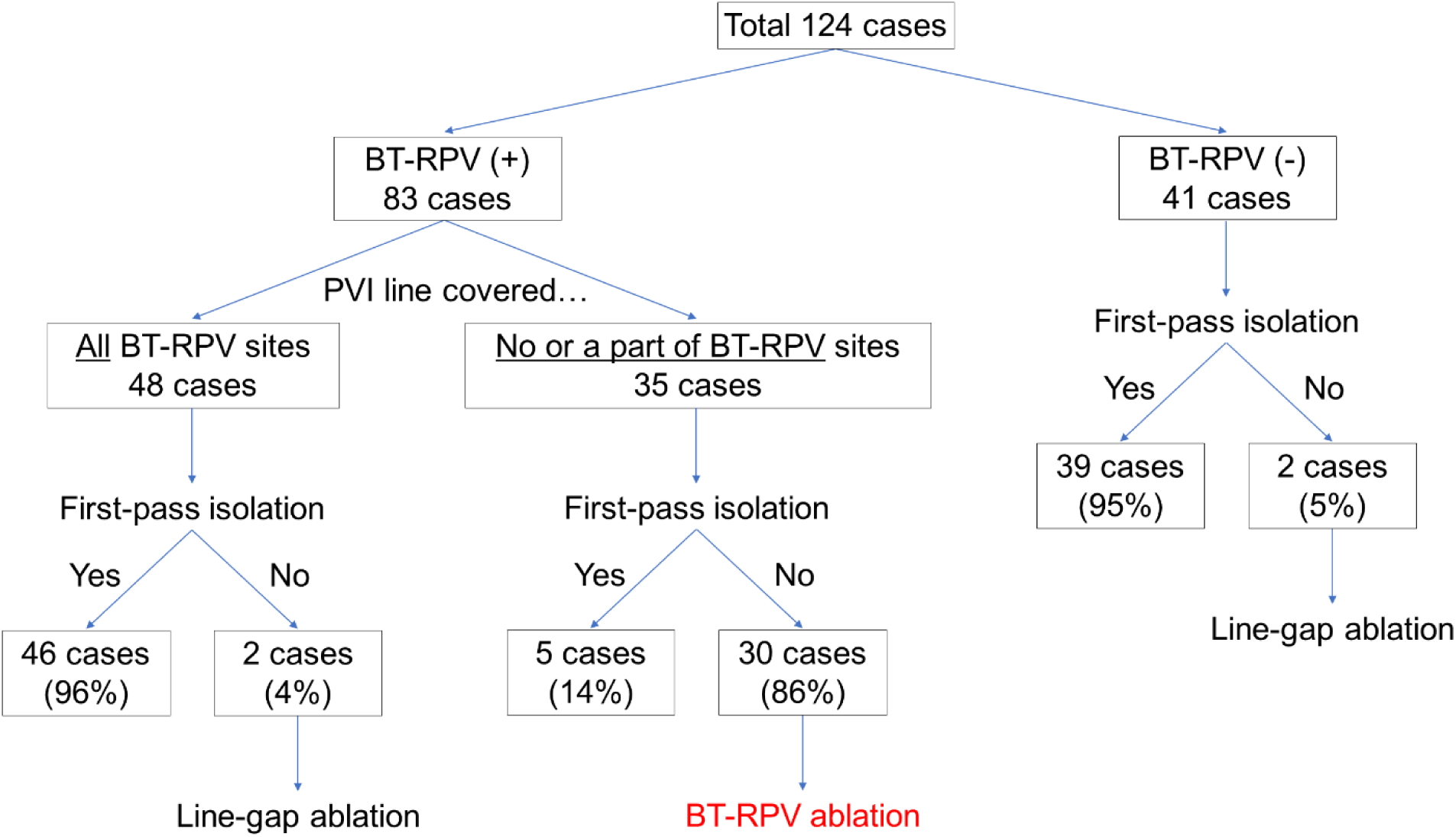
The outcome of the pulmonary vein isolation according to the presence or absence of the breakthrough of the activation into the right pulmonary vein antrum (BT-RPV). The details are described in the main manuscript. PVI indicates pulmonary vein isolation.

### Effects of ablation targeting the BT-RPV sites

The results of the ablation targeting the BT-RPV sites are summarized in Fig. 5. We targeted a total of 35 BT-RPV sites in 30 cases. The first BT-RPV ablation set successfully isolated the right PVs in 25 sites, while it led to delayed PV potentials in 8 sites and showed no effect in 2 sites. In the cases with 10 sites in which the initial BT-RPV ablation was insufficient for the PV isolation, additional ablation was performed to address the gaps in the PV isolation line, yet a complete PV isolation was not achieved in any of the cases. Among the 25 sites in which the PV isolation was initially successful, PV reconnections occurred at 5 sites. Consequently, a repeat BT-RPV ablation was performed for 15 sites in which the first ablation attempt did not completely isolate the right PVs, leading to a complete PV isolation in all those sites.

**Figure 5.**
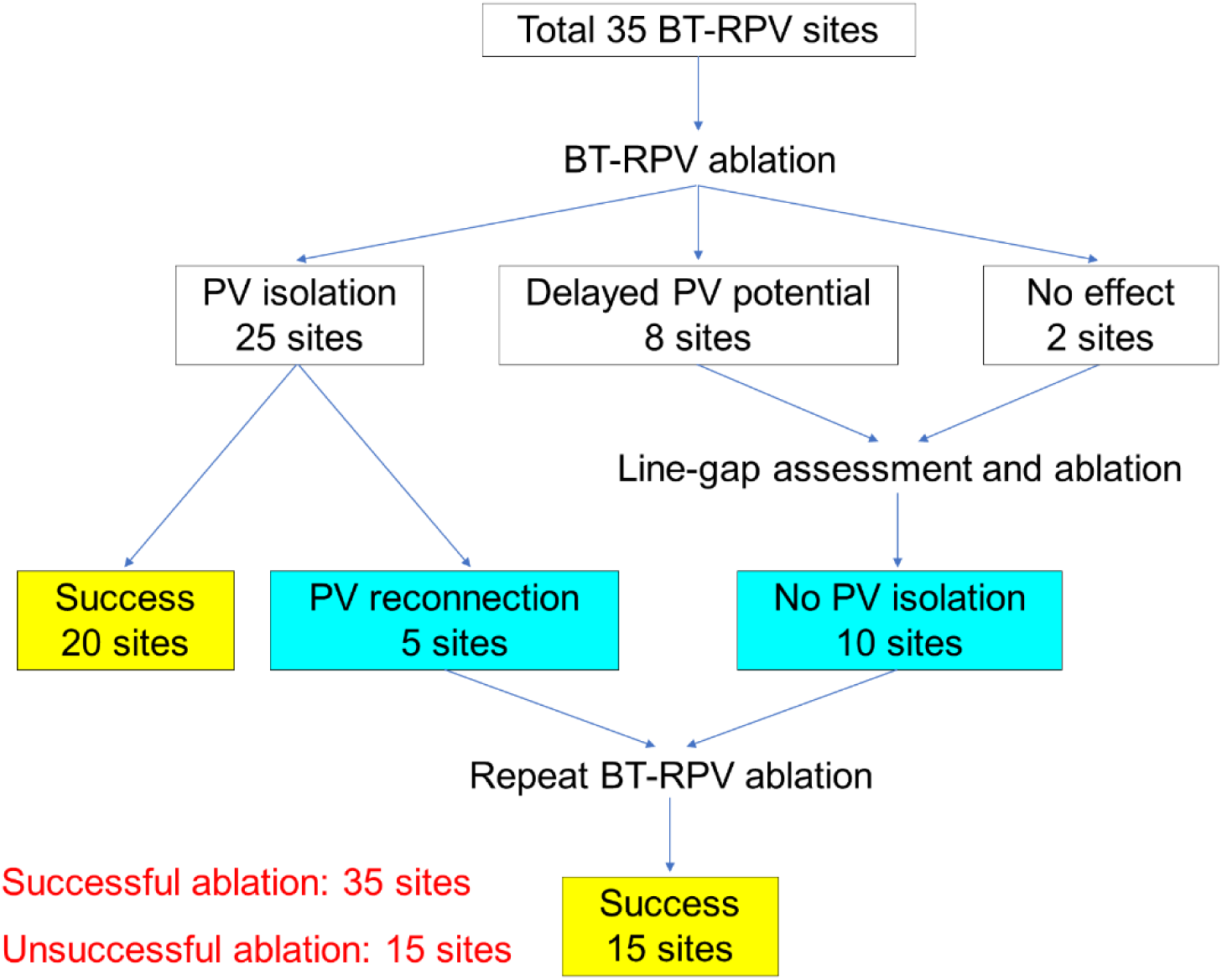
Outcomes of ablation targeting the breakthrough of the activation into the right pulmonary vein antrum (BT-RPV) for the pulmonary vein isolation. The details are described in the main manuscript. PV indicates pulmonary vein.

### Appropriate ablation lesion size for the BT-RPV ablation

The initial BT-RPV ablation was successful at 20 sites and unsuccessful at 15 sites, and a repeat BT-RPV ablation was successful at all 15 sites in which the initial ablation was unsuccessful. Therefore, we compared the properties of the BT-RPV ablation between the 35 sites with a successful BT-RPV ablation and 15 sites with an unsuccessful BT-RPV ablation.

The ablation lesion size was larger at the successful BT-RPV ablation sites than at the unsuccessful BT-RPV ablation sites (0.9 [0.6–1.2] cm^2^ vs. 0.6 [0.5–0.7] cm^2^, P = 0.010) (Fig. 6A). Furthermore, the ablation lesions at the successful BT-RPV ablation sites covered the area activated within a longer time after the emergence of the BT-RPV than that of the unsuccessful BT-RPV ablation sites (15 [14-16] ms vs. 10 [8–12] ms, P <0.001) (Fig. 7A). In the receiver-operating characteristic curve analysis, the ablation lesion size was determined as a predictor of a successful BT-RPV ablation with an area under the receiver curve of 0.731 (95% CI, 0.587–0.876) (Fig. 6B). A cutoff value of 0.7 cm^2^ had sensitivity, specificity, and positive and negative predictive values of 69%, 73%, 86%, and 50%, respectively. Furthermore, the time from the emergence of the BT-RPV to activate the area corresponding to the ablation lesion had a higher predictive accuracy with an area under the receiver curve of 0.963 (95% CI, 0.913–1.000) (Fig. 7B). A cutoff value of 14 ms had sensitivity, specificity, and positive and negative predictive values of 91%, 100%, 100%, and 83%, respectively.

**Figure 6.**
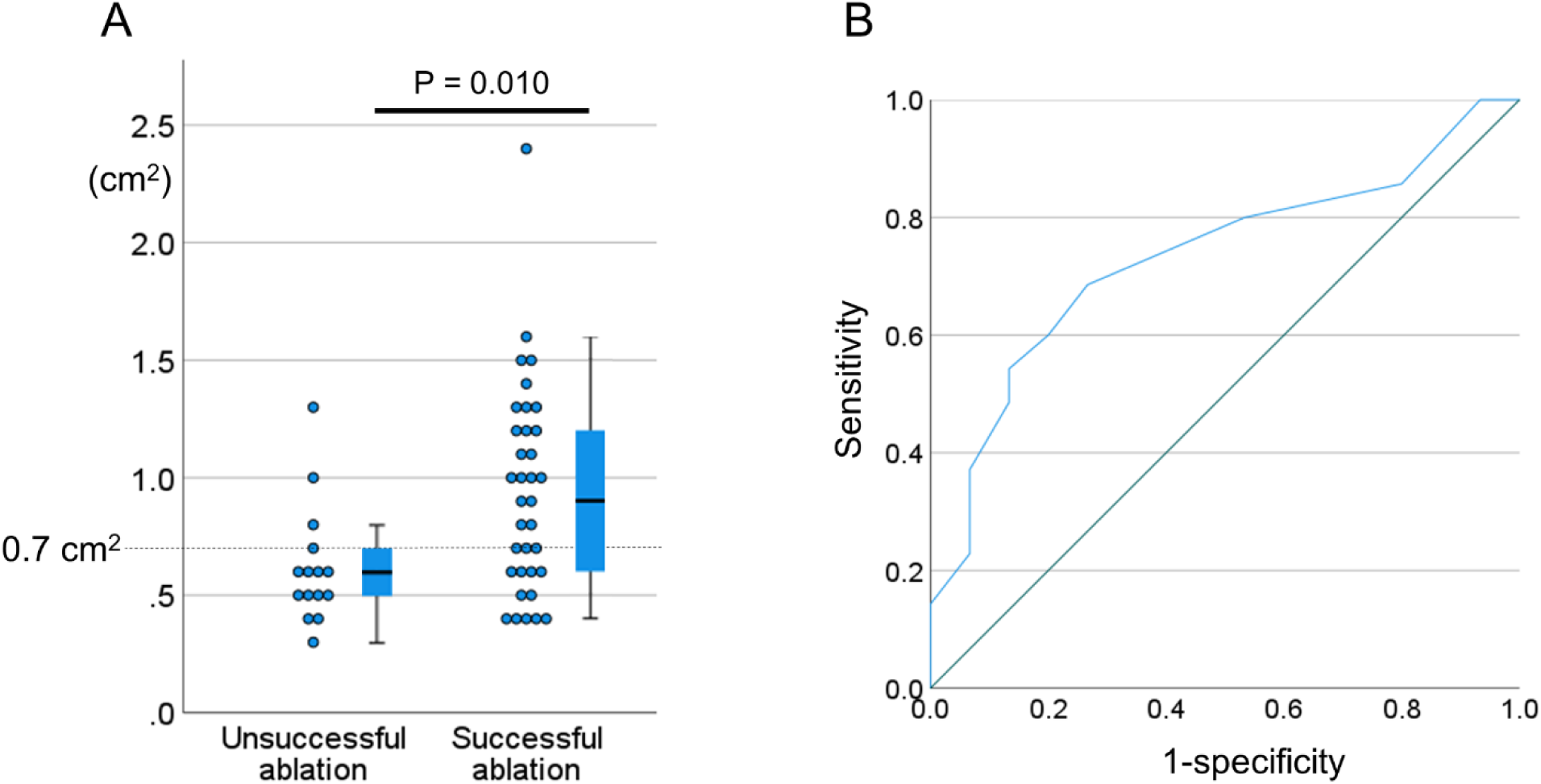
Comparison of the size of the ablation lesion and its predictive performance for a successful pulmonary vein isolation. (A) A box-whisker plot of the lesion size of the ablation targeting the breakthrough of the activation into the right pulmonary vein antrum (BT-RPV) at the successful and unsuccessful ablation sites. (B) A receiver operating characteristic curve for the prediction of a successful pulmonary vein isolation based on the size of the BT-RPV ablation lesion.

**Figure 7.**
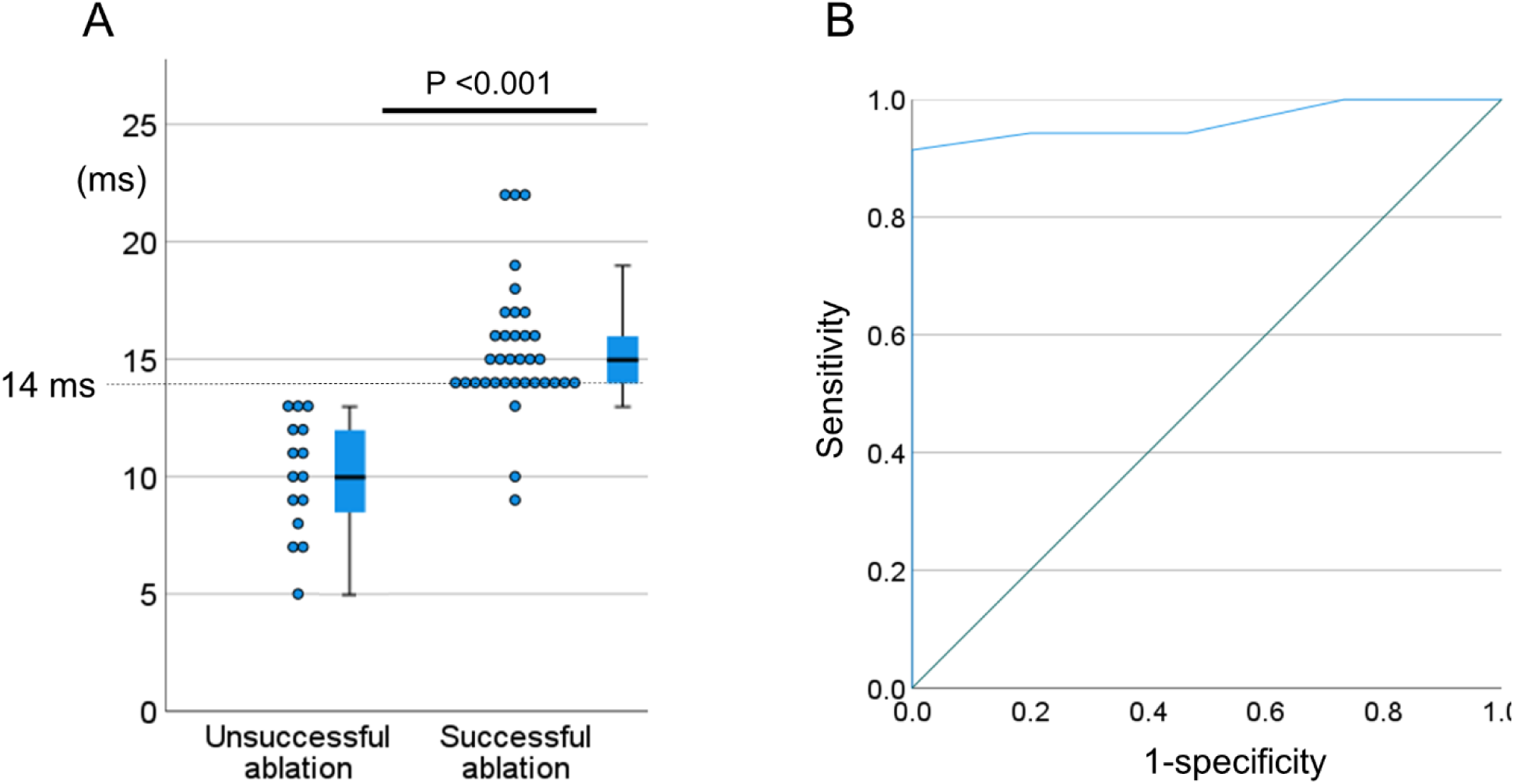
Comparison of the time needed to activate the area corresponding to the ablation lesion and its predictive performance for a successful pulmonary vein isolation. (A) A box-whisker plot of the time from the emergence of the breakthrough of the activation into the right pulmonary vein antrum (BT-RPV) to activating the area corresponding to the ablation lesion at the successful and unsuccessful ablation sites. (B) A receiver operating characteristic curve for the prediction of a successful pulmonary vein isolation based on the time needed to activate the area corresponding to the ablation lesion.

### Complications and AF recurrence

No major complications, such as strokes, cardiac tamponade, or esophageal damage were observed perioperatively. During a 14 [11–18] month follow-up, AF recurrence was noted in 5 cases with BT-RPVs and 1 case without. The AF-free survival rates were 94.0% in the patients with BT-RPVs and 97.6% in those without. The log-rank test showed no significant difference in the AF-free survival between the patients with and without BT-RPVs (P = 0.386). A repeat catheter ablation was performed in 2 cases with BT-RPVs, exhibiting no right PV reconnections. In contrast, 1 case without a BT-RPV underwent the procedure, revealing a right PV reconnection due to a gap in the PV isolation line.

## DISCUSSION

To the best of our knowledge, this study is the first to assess the effectiveness of the ablation approach targeting BT-RPV sites detected before the PV isolation. Its main findings were as follows:

1. A BT-RPV was observed in 67% of patients with AF, including 14% of patients who had two distinct BT-RPV sites.
2. BT-RPV sites were most frequently located on the anterior part of the carina (45% of BT-RPV sites), followed by the anterior part of the right inferior PV antrum (24%), and the middle part of the carina (17%).
3. In patients with BT-RPVs, a right PV isolation can be challenging. However, the success rate of the first-pass ablation remained high at 96% as long as the isolation line covered all BT-RPV sites. That rate was comparable to the 95% success rate in the patients without BT-RPVs.
4. Although ablation targeting BT-RPV sites is effective for a right PV isolation, a certain area of the ablation lesion is needed for a durable PV isolation. Ablating the area that is activated within 14 ms after the emergence of the BT-RPV provided a high success rate of the PV isolation.
5. The ablation strategy targeting the BT-RPV identified prior to the PV isolation resulted in comparable AF-free survival rates in patients with a BT-RPV as compared to those without. That outcome was attributed to the durable isolation of the right PVs.

### Anatomical substrates for epicardial conduction to the right PV antrum

There are two possible substrates for the epicardial pathway to the right PV antrum. One is the connection with the right atrium through intercaval muscular fibers. In some hearts, there are broad muscular bridges across the posteroinferior interatrial groove, joining the left atrial musculature to the intercaval area of the right atrium and to the insertion of the inferior caval vein.^4,5^ Furthermore, small muscle bridges are observed in some hearts connecting the muscular sleeves of the right PVs to the right atrium and the superior vena cava.^4,5^ Those muscular fibers may form electrical conduction pathways connecting the anterior part of the right PV antrum to the posterior wall of the right atrium and the superior vena cava.^6–9^ The other possible substrate for the epicardial pathway is the rightward branch of the septopulmonary bundle. The septopulmonary bundle is the epicardial muscular fiber emerging from the interatrial groove and running through the posterior wall of the left atrium.^4,5,11,12^ On the posterior wall of the left atrium, the septopulmonary bundle often bifurcates to become 2 oblique branches, and the rightward branch turns into the posterior septal raphe. Therefore, this rightward branch of the septopulmonary bundle may electrically connect the posterior part of right PV antrum to the left atrium. A previous study^8^ suggested that this type of epicardial connection is less frequent than that through the muscular fibers from the right atrium, which is consistent with our result that BT-RPVs were less frequent in the posterior part of the right PV antrum.

### Impact of epicardial connections to right PV antrum on the ablation outcome

Epicardial connections have been previously identified as factors that impede a successful PV isolation. The cases featuring epicardial connections frequently require additional ablation at the carina, which subsequently needs a longer radiofrequency application time.^6^ Moreover, the presence of an epicardial connection reduces the chance of a successful PV isolation and increases the recurrence of atrial arrhythmias.^8^ In contrast to the previous study,^8^ our research observed high success rates of a PV isolation and low AF recurrence rates, regardless of the presence or absence of BT-RPVs. This discrepancy can be attributed to the fact that in our study, the BT-RPV ablation was performed over a relatively wider area.

In our research, we noted a much higher prevalence of epicardial connections (67%) than that observed in the earlier studies 7.1–18.0%).^6,8^ This variance could partly be ascribed to the differences in the size of the PV isolation area. More crucially, epicardial connections were assessed before the PV isolation in our study, whereas the epicardial connections were assessed after the PV isolation in the previous study.^8^ Therefore, it is hypothesized that a substantial number of epicardial connection sites were inadvertently ablated during the process of the WACA. Interestingly, the PV isolation was achieved without a BT-RPV ablation in 5 cases in whom the detected BT-RPV sites were not covered by the PV isolation line. That finding suggested that the heat from the ablation for the PV isolation remotely modified the epicardial pathways in a part of the cases.

### Clinical implications

There were two advantages of assessing the BT-RPV before the PV isolation. First, after the PV isolation, the potentials inside the isolation line became smaller as compared to those outside the line. That could make it challenging to assess the BT-RPV sites due to the inappropriate annotations of the far-field signals from the atrial muscle outside the isolation line. That issue did not exist when the mapping was performed before the PV isolation. On the other hand, when evaluating the BT-PRVs prior to the PV isolation, the electrical activation via other interatrial conduction pathways could obscure the BT-RPV. However, activation via intercaval muscular fibers typically precedes that of other pathways because those fibers enable direct conduction from the right atrium, in contrast to other pathways that involve excitation through the left atrium. Second, by evaluating the BT-RPV sites before the PV isolation and designing the PV isolation line to pass through them, the success rate of the first-pass ablation for the PV isolation could be increased, thus avoiding excessive ablation.

Even when BT-RPVs cannot be covered by the PV isolation line, direct ablation of the BT-RPV sites can block the conduction due to epicardial connections. It is important to note that the connections of epicardial pathways are broad; consequently, an extensive area must be ablated during the BT-RPV ablation to achieve a durable PV isolation. Since the extent of epicardial pathway connections vary among individuals, the lesion size by itself is not a sufficient marker for determining the appropriate ablation area. Considering the simultaneous excitation of the connection sites of an epicardial pathway, it is essential to target the area activated within a certain timeframe (14 ms in this study) after the emergence of the BT-RPV.

### Study limitations

This study had several limitations. First, this study was conducted with a relatively small number of cases from a single center. Second, in this study, an evaluation of the epicardial connections outside the PV line using PV pacing was not conducted, thus the origins of each pathway were not clear. Third, although the study was carried out prospectively, the appropriate ablation lesion area was analyzed retrospectively. Therefore, the criteria for the appropriate ablation lesion demonstrated in this study need to be validated prospectively. Last, the recurrence of atrial arrhythmias was diagnosed by clinical interviews and clinical examinations using a surface 12-lead electrocardiogram and Holter monitor, which is a method that is known to underestimate the prevalence of atrial arrhythmias.^14^

## CONCLUSIONS

BT-RPVs were observed in 67% of the patients with AF. Although BT-RPVs can complicate the right PV isolation, the success rate of a first-pass ablation remained high, provided the isolation line covered all the BT-RPV sites. Ablation targeting BT-RPV sites proves effective in cases where WACA fails to achieve a right PV isolation. However, to ensure a durable PV isolation, it is crucial to create a sufficiently extensive ablation lesion due to the broad connections of the epicardial pathways. Notably, ablating the area activated within 14 ms following the emergence of the BT-RPV yielded a high success rate of the PV isolation. Furthermore, implementing this ablation strategy was associated with a high rate of AF-free survival.

## Data Availability

The data that support the findings of this study are available from the corresponding author upon reasonable request.

## Non-standard Abbreviations and Acronyms

AF: atrial fibrillation
BT-RPV: breakthrough of the activation into the right pulmonary vein antrum
PV: pulmonary vein
WACA: wide antral circumferential ablation

## Acknowledgements

The authors would like to thank Mr. John Martin for the English language review.

